# Training and testing a CNN-based engine for brain MRI scan classification and segmentation

**DOI:** 10.1101/2023.07.03.23292122

**Authors:** Kunaal Dhawan, Utkarsh Agrawal

## Abstract

Biomedical imaging data (from X-rays, CT scans, MRIs) is a rich and crucial source of information for diagnosing and treating patients, yet the large volume of visual data generated can be challenging even for the most experienced clinical professionals to handle and use for diagnosis with high accuracy. With the traditional diagnosis approaches, patients usually face high costs too. Sometimes, abnormalities remain unspotted, causing delays in intervention which can be the difference between saving and losing patient lives. Most traditional radiomics studies use hand-crafted feature extraction techniques, such as texture analysis, followed by conventional machine learning classifiers, such as random forests and support vector machines (SVMs). In this paper, we establish that X-ray, MRI or CT-image classification using convolutional neural networks (CNNs) could be an efficient, cost-effective and fast approach for diagnosis and interpretation. The research is focused on training a CNN algorithm to develop diagnostic analysis power using 250 brain MRI images and then testing the accuracy and predictive power of the developed CNN algorithm on 5 images. We have shown the superiority of CNN models in image classification compared to traditional ML Classifiers that used an extensive ablation study. We have then compared a selected set of existing models with the new model we have built based on the EfficientNet v2-S architecture with a high classification accuracy of 98% – showing a high predictive power to provide various differential diagnoses from the brain MRI scans.

## 1. Introduction

Magnetic resonance imaging (MRI) is used less commonly than the plain X-rays and computer tomography (CT) scans, yet it is best for viewing and interpreting soft tissues such as fat, water, muscle, and cerebrospinal fluid (CSF) inside the brain for neurological and musculoskeletal pathology. Based on a study [1], nearly 82% of the brain MRI scans come out normal, while the rest require further diagnoses and clinical correlation. While only 1.2% of the brain MRIs require urgent or immediate referral, sifting through voluminous sets of images and arriving at the ones that show abnormalities needing further assessment is very onerous.

While imaging data from brain MRI scans is an incredibly useful and rich source of information about patients for neurological conditions, it is often one of the most complex one. Moreover, with the traditional diagnosis techniques, patients usually face high costs and delays in care delivery as sometimes it may even take months before a life-threatening condition is discovered. Sometimes, even the slightest abnormalities remain unspotted, which can be the difference between saving and losing the life of a patient.

The artificial intelligence (AI) based algorithms can scan through the image to recognize anomalies like lesions, tumors, musculoskeletal injuries, and so forth. AI-algorithms can spot even the slightest of abnormalities invisible to the human eye, making them an ultimate tool for improvised and effective medical diagnosis. The use of AI-based algorithms serve a two-fold purpose by giving additional resources in the hands of the patients to get their biomedical images diagnosed with just a click of a button and drastically save their time and resources while also providing a second opinion to radiologists and their staff.

To tackle these challenges, we have developed an AI-based MRI scanning engine that can help identify features in MRI biomedical images quickly and precisely, by simply uploading the MRI scan images in the application that has the diagnostic engine. This research is focused on training a convolutional neural network (CNN) algorithm to develop diagnostic analysis power using 250 brain MRI images and then testing the accuracy and predictive power of the developed CNN algorithm on 5 images. An EfficientNet v2 [2] based CNN algorithm has been developed in this study. To demonstrate the power of CNN algorithms in extracting contextual information as well as spatial features from an input image, we also compare the predictive performance of traditional statistical ML algorithms on the same dataset with our proposed CNN-based engine. To build these models, we have designed and deployed the neural networks on the Kaggle Cloud ML Engine that supports Tensorflow API.

## 2. Brief literature review

It is well acknowledged that computer vision based on convolutional neural networks (CNNs) have enabled practitioners in various fields to innovate on applications such as face recognition, autonomous vehicles, self-service supermarkets, and intelligent medical treatments [3]. CNN is a type of deep learning model for processing data that has a grid pattern, such as images, which is inspired by the organization of visual cortex and designed to adaptively learn spatial hierarchies of features, from low-level to high-level patterns. CNN is a mathematical construct that is typically composed of three layers (or building blocks): convolution layer, pooling layer, and fully connected layer. The first two, convolution and pooling layers, perform feature extraction, while the third, a fully connected layer, maps the extracted features on to final output, such as classification. The convolution layer plays a key role in CNN, which is composed of a stack of mathematical operations. In digital images, pixel values are stored in a two-dimensional (2D) grid, i.e., an array of numbers and a small grid of parameters called kernel. A kernel is a filter in the form of an optimizable feature extractor that is applied at each image position, which makes CNNs highly efficient for image processing, since a feature may occur anywhere in the image. As one layer feeds its output into the next layer, extracted features can hierarchically and progressively become more complex. The process of optimizing parameters such as kernels is called training.

The object detection is at the core of using CNN for various predictive applications and any engine built using CNN not only has to recognize the category of images being interpreted but also the segmentation of the image at the next level with the right set of classifiers. Most traditional radiomics studies use hand-crafted feature extraction techniques, such as texture analysis, followed by conventional machine learning classifiers, such as random forests and support vector machines [4]. There are several differences to note between these traditional methods and CNN. Firstly, CNN does not require hand-crafted feature extraction. Secondly, CNN architectures do not necessarily require segmentation of tumors or organs by human experts. Thirdly, CNN is far more data hungry because of its millions of learnable parameters to estimate which makes it computationally more expensive, resulting in requiring graphical processing units (GPUs) for model training.

The recent applications of CNN in radiology are divided into four categories: classification, segmentation, detection, and others. In medical image analysis, classification with deep learning utilizes target lesions depicted in medical images, and these lesions are classified into two or more classes, for example, benign or malignant. For segmentation, one of the common ways is to use a CNN classifier for calculating the probability of the image being an organ or anatomical structure. In this approach, the segmentation process is divided into two steps; the first step is the construction of the probability map of the organ or anatomical structure using CNN and image patches, and the second is a refinement step where the database of images and the probability map are utilized. Finally, the most important step for radiologists is to detect abnormalities within medical images. Abnormalities can be rare and hard to spot, and they must be detected amongst a majority of normal cases.

The research study by Hasnain Ali Shah et. al. [5] compared the performances and efficiencies of six CNN architectures: VGG16, GoogLeNet, InceptionResNetV2, Xception, ResNet50, and EfficientNet-B0. Each deep CNN in the study employed the same set of parameters and features that varied according to the depth of the convolution layer and the fully connected layers. All models showed a minimal error gap at the end of each phase, except for InceptionResNetV2, which had a slight overfitting problem at the beginning. All the other models showed a very stable minimization of loss. In the first study, EfficientNet-B0, a deep neural network developed by Google AI was employed to investigate the transfer learning approach for detecting the brain tumors in MRI scans. The proposed fine-tuned EfficientNet-B0 network achieved the highest (98.87%) accuracy on the validation data by outperforming the other networks discussed below.

## 3. Methodology

The proposed research performs the task of identifying whether an input brain MRI scan contains a tumor or not. This is a classical binary classification problem which adopts the workflow shown in Figure 1. From a given dataset containing brain MRI scans of various patients, we first perform a few pre-processing steps to ensure the images are in the requisite format for training the CNN architecture. We apply normalization by thresholding and dilating each image to remove excess noise. We also crop each image with dynamic coordinates so that excess portion outside the brain’s cross-section is removed, thus, further de-noising the image. Finally, we reshape each image to fit the dimensions 150 × 150 as a standard across the entire dataset.

**Figure 1.**
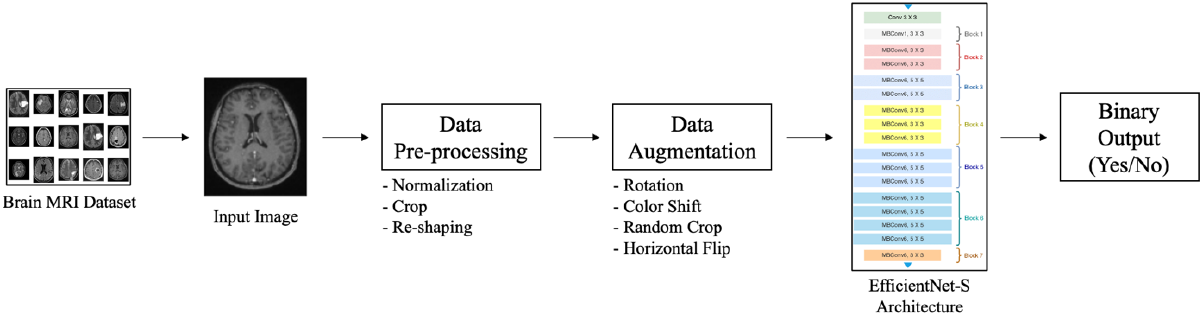
Workflow of the proposed algorithm.

After pre-processing, we augment the dataset through rotation of axes, horizontal flips, color shifting and random cropping, in order to increase the size of our database, thus, ensuring the learning algorithm has no bias to the 250 images present within the dataset.

Once we have the pre-processed and augmented set of images, we train the EfficientNet v2-S architecture configuration with an input of shape 150 × 150. EfficientNet v2-S is a version of EfficientNet v2 architecture which is pre-trained on ImageNet [6] dataset and is widely used in performing various image classification tasks achieving state-of-the-art performance. The output probabilities from the network are used to assign a class to the input image, which in turn is used to identify whether the MRI scan is a tumor or not.

### 3.1. Dataset

We use a dataset titled ‘Brain MRI Images for Brain Tumor Detection’ [7], available on Kaggle as an opensource database of 253 Brain MRI scans belonging to two classes: containing a tumor or not. Out of 253 raw images, a total of 155 images show a tumor and 98 images are benign.

### 3.2. Data Pre-processing and Augmentation

MRI scans can falsely induce intensity averages due to various distortions introduced to the system while scanning the subject. This condition may lead to the algorithm falsely identifying a benign MRI scan containing tumor, therefore, the pre-processing stage is extremely important. We perform normalization of the entire image which in essence is a function to reduce the intensity values to come within a range suitable for the CNN to learn the spatial characteristics of the MRI scan. This step sets the mean intensity close to 0 and standard deviation close to 1. Since our network takes an input of shape 150 × 150, we re-shape each image to this dimension before feeding it to our CNN.

Since the dataset is a small database of MRI scans, we also perform data augmentation to increase the amount of training images fed to the CNN model for optimizing the cost function. This process generates additional training data based on the original scans through a series of transformations which preserve the true nature of each scan and adds some variance within the dataset. Each image in the training set is rotated within a range of 20 degrees randomly, either clockwise or anti-clockwise. We also shift the height and width dimensions by a factor of 20%. Similarly, we apply shear and zoom transformations which are 0.2 times the original dimensions. Finally, we also flip each image across the horizontal axis, thus, preserving their spatial references. The pre-processed and augmented set of training images are then fed to the CNN network whose architecture is described in the next sub-section.

### 3.3. EfficientNet v2-S Architecture

Since CNNs can learn features within training images and classify appropriately using both visual and spatial information, they are a suitable choice for our proposed architecture instead of using filters to perform image classification. We employ the technique of transfer learning where the network is first pretrained on a large-scale dataset, typically, ImageNet [6], and the learned weights of the parameters are used as a starting point to optimize the cost function generated using the given small dataset. This works under the hypothesis that spatial features like edges, curves, and so forth, that are learned using a large-scale dataset can be translated to smaller-scale datasets. We leverage this property to effectively deploy the EfficientNet v2-S architecture for classifying the images in our database of Brain MRI scans.

EfficientNet [8] is a family of CNNs which employ various scaling methodologies to uniformly re-scale each dimension of the input using a dynamic coefficient called compound coefficient. Usually, in a generic CNN, these dimensions (height, width, number of channels) are randomly scaled with factors learned during the training procedure. EfficientNet makes this process uniform by fixing the scaling coefficient using methods described in Figure 2. We can intuitively verify this by assuming that if the resolution of input image is considerably high, the number of layers required within the network would also be larger to increase the receptive field. This also means that we require more and more channels to learn the patterns existing within the receptive field. The base network is designed using inverted bottleneck residuals, squeeze and excitation blocks proposed in MobileNet v2 [9].

**Figure 2.**
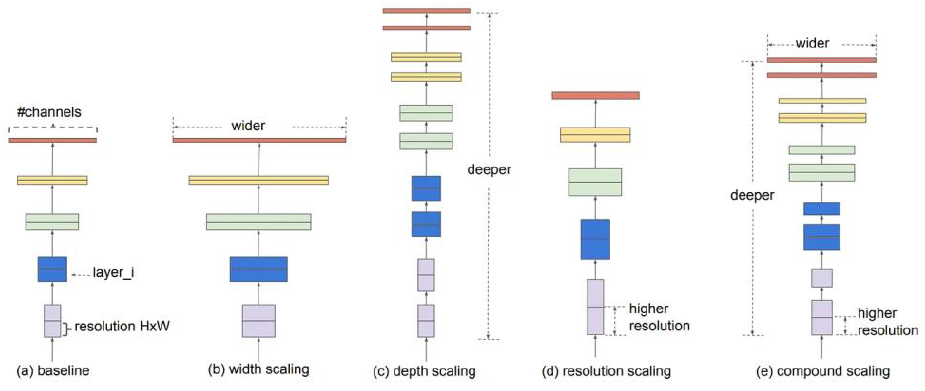
Scaling techniques used in EfficientNet [8].

EfficientNet v2 architecture improvises upon EfficientNet in terms of parameter efficiency and time required by the cost function to reach global minimum. We leverage the S configuration of EfficientNet v2 architecture which is the most light-weight configuration containing a total of 22M training parameters requiring a total of 8.8B FLOPs. This configuration ensures that we can design and deploy our model on a computational platform with minimal requirements in terms of both memory and efficiency. Moreover, the EfficientNet v2-S configuration takes around 7.1 hours to train on the ImageNet dataset which is relatively quick in comparison to other state-of-the-art classifiers like Vision Transformer ViT-L [10], which takes around 172 hours for convergence.

Table I summarizes the architecture of EfficientNet v2-S used in our research work. EfficientNet v2 accepts a dynamic input size, therefore, we set it to 150 × 150 based on our dataset. The input is relayed to the first stage containing a 3×3 Convolution block with stride 2 and total of 24 channels. The feature maps generated using this single Convolution block are fed into 10 layers of Fused-MBConv blocks of kernel size 3×3 each. These blocks have varying strides of 1, 2, and 3 across 3 stages, and contain 24, 48, and 64 channels respectively. The next 3 stages contain MBConv blocks each, with large number of channels. There is a total of 30 such layers followed by a single 1×1 Convolution, Pooling, and Fully connected layer. The input number of channels for this final stage is set to 1280. Since we are performing transfer learning, we remove the top layer of EfficientNet v2-S and add a few custom layers. First, we flatten the output to a total of 32000 pixels and apply a Dense block of dimension 1024. To regularize the training process and re-parametrize the input, we add a Dropout layer with a threshold probability of 0.2. Finally, we attach an output layer with Sigmoid activation to output the probability having a single dimension as it is a binary classification problem.

**Table I.** EfficientNet v2-S architecture

| Stage | Block | Channels | Layers |
| --- | --- | --- | --- |
| **0** | 3x3 Conv | 24 | 1 |
| **1** | 3x3 Fused-MBConv1 | 24 | 2 |
| **2** | 3x3 Fused-MBConv4 | 48 | 4 |
| **3** | 3x3 Fused-MBConv4 | 64 | 4 |
| **4** | 3x3 MBConv4 | 128 | 6 |
| **5** | 3x3 MBConv6 | 160 | 9 |
| **6** | 3x3 MBConv46 | 256 | 15 |
| **7** | 1x1 Conv + Pooling + FC | 1280 | 1 |
| **8** | Flatten | 32000 | 1 |
| **9** | Dense | 1024 | 1 |
| **10** | Dropout | 1024 | 1 |
| **11** | Dense | 1 | 1 |

### 3.4. Implementation steps used

To train the architecture deployed in our work, we split the dataset into 3 sets used for training, validation and testing with 161, 41, and 51 images each respectively. The pretrained weights of EfficientNet v2-S on ImageNet were pre-downloaded using TensorFlow library and the network was further finetuned across all the layers from start to end after addition of stages 8 to 11. The CNN was trained for a total of 20 epochs with a batch size of 16 images. We used binary cross entropy loss as our cost function and Adam optimizer with a learning rate of 1e-4. The end-to-end design and development for this research has been done in Jupyter Notebook hosted on Kaggle Cloud ML Engine.

## 4. Experimental results

We present the results from the quantitative analysis performed on the test set of the given brain MRI database using three key metrics: total number of trainable parameters, time taken for inferencing per step, and classification accuracy. Total number of trainable parameters are simply the weights whose values are learned and updated during the optimization procedure of the CNN. The key reason for optimizing for the number of weights is due to the fact that the overall size of the model is directly proportional to the total number of weights it has to update as these are saved as floating points in the computer memory. Finally, to compare the efficacy of tumor classification, we simply compute the classification accuracy using Equation 1.

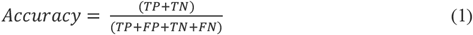

Here, TP denotes the total number of true positives detected by the CNN, i.e., images classified as tumorous which were truly malignant. TN is defined as true negatives, which is the total number of images detected benign which were truly benign. Similarly, FP (false positives) and FN (false negatives) are incorrectly classified images with total number of tumorous detections which were truly benign and total number of benign detections which were truly malignant, respectively. We perform quantitative analysis while comparing our key CNN EfficientNet v2-S, with various state-of-the-art image classifiers, namely: Inception ResNet v2 [11], Inception v3 [12], MobileNet v2 [9], ResNet-101 v2 [14], and Xception [15].

These classifiers demonstrate extremely strong performance when it comes to image classification, specially, on the large-scale ImageNet dataset. Therefore, we compare the performance with our proposed methodology based on EfficientNet v2-S to understand the pros and cons of our CNN-model architecture.

Table II summarizes the quantitative analysis performed in this research on various CNN architectures along with our proposed methodology denoted by ‘EfficientNet v2-S’. It is evident that our architecture choice (shown as “recommended”) outperforms various other state-of-the-art image classifiers in the transfer learning task of identifying tumor within Brain MRI scans by achieving a high classification accuracy of 98.03%. Inception ResNet v2 achieves performance closer to our architecture, however, takes more size and runtime to compute the classification decision. EfficientNet v2-S requires 22.6% less memory in terms of trainable parameters and runs faster by 88ms per step in comparison to Inception ResNet v2. MobileNet v2, which is one of the most efficient and light-weight CNN, performs the classification task on our dataset by using only 34.99M parameters and 45ms, however, fails to achieve high classification accuracy as the model fails to generalize on our test set. Therefore, our proposed approach detects tumors in a set of unseen images with high accuracy. least computational requirement, and faster runtime, making it a suitable CNN architecture to be deployed on the mobile systems.

**Table II.**

| **Algorithm** | **# Params ↓** | **Inference Time / Step ↓** | **Accuracy ↑** |
| --- | --- | --- | --- |
| **Inception ResNet v2** | 68.43M | 325ms | 92.15% |
| **Inception v3** | 40.64M | 125ms | 86.27% |
| **MobileNet v2** | **34.99M** | **45ms** | 82.35% |
| **ResNet-101 v2** | 94.95M | 447ms | 86.27% |
| **Xception** | 73.23M | 278ms | 86.20% |
| **EfficientNet v2-S**<br>**(Recommended)** | 52.94M | 237ms | **98.03%** |

We also plot the training as well as validation curves for the CNN architectures of our choice in order to understand the optimization process and how well the architecture generalizes to the unseen validation set. Figure 4 showcases the change in training accuracy per epoch along with the reduction in objective function’s cost or loss across each epoch. Similar curves are depicted in Figure 5 for the validation set.

**Figure 4.**
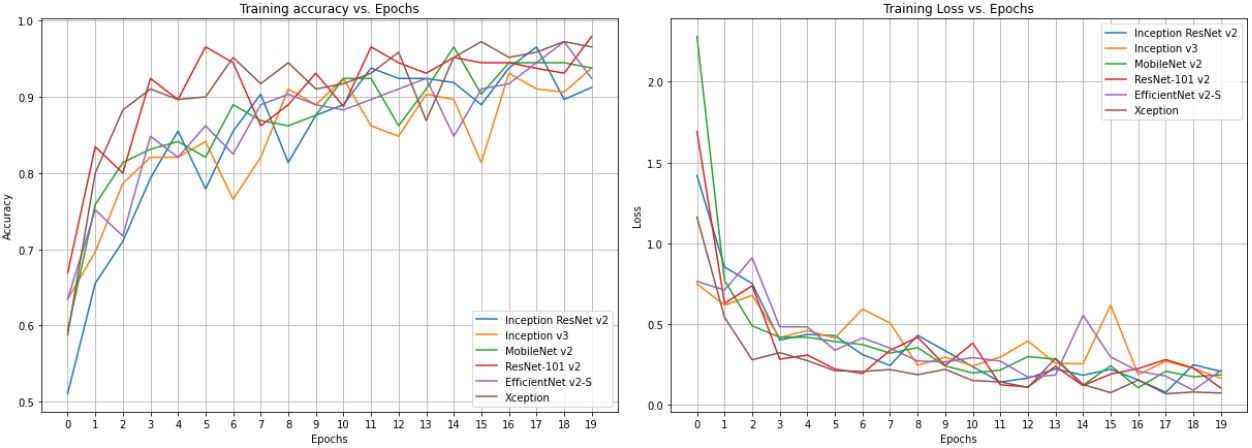
Training curves.

**Figure 5.**
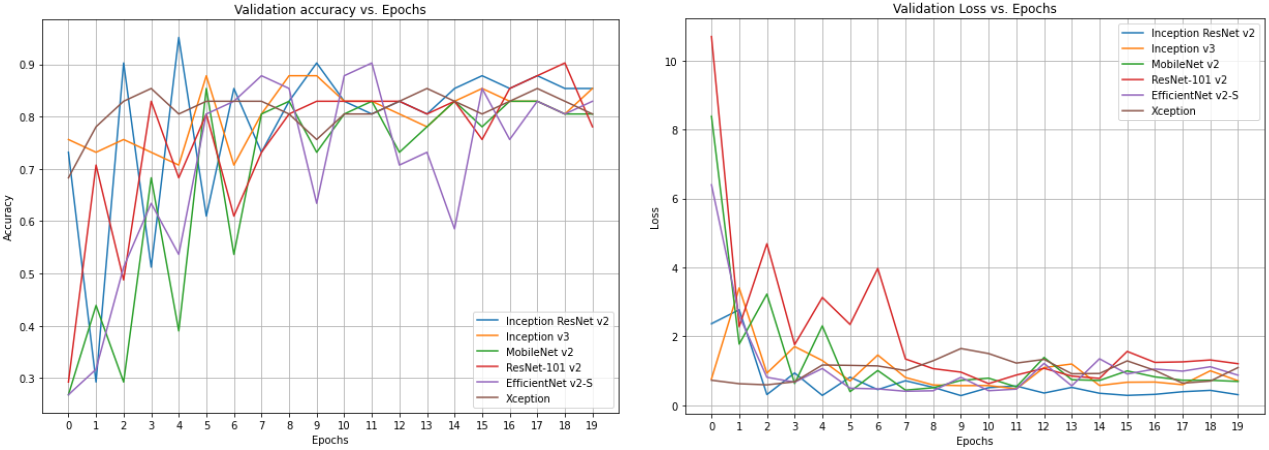
Validation curves.

To benchmark the advantage of feeding images directly into a CNN instead of using traditional filter-based feature classification using statistical ML classifiers, we compare the predictive performance on the same dataset by using 3 powerful algorithms: K-Nearest Neighbors (K-NN), Decision Trees and SVM. For this, we create a separate pipeline which pre-processes the dataset and passes it on through several filters to extract some features from the image data. First filter called Gray-Level Co-Occurrence Matrix (GLCM) examines the existing textures within the image by monitoring the spatial relational of each pixel. It basically computes the count of pair of pixels containing specific values and a set spatial context, generating a matrix of feature transformations. We also use a technique called Local Binary Pattern Feature Detection (LBPFD) [15] which describes the texture information of each image in the form of threshold-based neighboring values of the current pixel. With this approach, we are able to capture spatial patterns within the grayscale image with the help of the contrast. Once we generate the features, we simply feed them into statistical classifiers to compute classification accuracy on the test set.

Figure 6 compares the classification accuracy of CNN-based methods and ML classifiers. Our recommended approach based on EfficientNet v2-S clearly outperforms ML Classifiers by achieving 98.03% classification accuracy. ML classifiers fail to achieve comparable accuracy to other CNN-based architectures. This indicates that filters fail to capture the spatial context and representation-based features within the learning task of brain tumor detection, and therefore, CNNs are superior in performance to ML classifiers.

**Figure 6.**
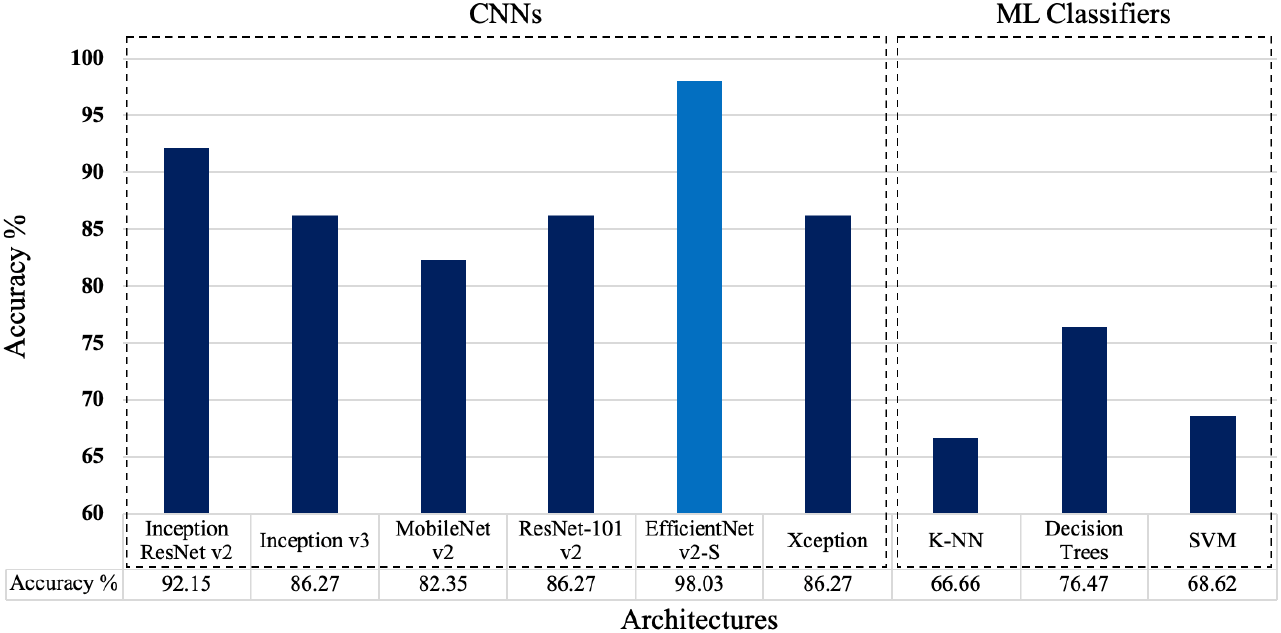
Accuracy comparison OF CNN architectures with ML classifiers.

## 5. Conclusion

We leveraged the rich information present in the biomedical imaging data of Brain MRI scans to identify whether a given Brain MRI has tumor or not. Our research focused on benchmarking various CNN algorithms to analyze the visual features in MRI images and learn patterns to diagnose a patient efficiently and accurately. We proposed an end-to-end pipeline based on state-of-the-art EfficientNet v2-S architecture to classify a given brain MRI scan as benign or malignant. Our architecture achieved a high classification accuracy of 98.03%, while requiring minimal memory based on 52.94M trainable parameters. The proposed architecture performs inferences in 237ms per step, which is fast in comparison to state-of-the-art image classifiers like Inception ResNet v2, Xception, etc. as depicted through our quantitative evaluation. We also showcased the superiority of CNN models in image classification when compared to traditional ML Classifiers by performing an extensive ablation study. In conclusion, we studied the design of various CNNs and ML classifiers to propose a highly efficient and accurate architecture to identify brain tumors.

## 6. Limitations and ideas for future research

Though statistically significant, the training set of ∼250 images and the testing set of 5 images could be further enhanced, especially we could add more images for testing the predictive accuracy. In addition, the input images resized to 150 × 150 could be augmented further in the subsequent research to achieve even higher accuracies for a few of the architectures studied. Finally, a test-run along with a trained physician or a radiologist could further help get to qualitative insights on the predictive accuracy, along with any obvious ‘misses’ that the model may have done.

Towards future research, we could explore the application of GenAI (generative AI) through using Vision-Language foundation models to the brain MRI dataset to be able to create and train an engine/capability to re-create the key aspects of abnormalities detected from the CNN-based diagnosis s into reliable visual images. This has been done for the Chest X-ray generation [16].

## Data Availability

All data as well as the full source code used in creating and training the new CNN-based engine presented in the present study are available upon reasonable request to the authors.

https://www.kaggle.com/datasets/navoneel/brain-mri-images-for-brain-tumor-detection?datasetId=165566

## 7. Acknowledgements

We are thankful to Mr. Siddharth Nijhawan (M.S., Columbia University, New York) for providing with guidance on analytics and comparative assessment across various CNN models throughout the project.

## References

[1] Incidental findings on brain magnetic resonance imaging from 1000 asymptomatic volunteers; G. L. Katz man et al. JAMA, 1999

[2] The basics of MRI interpretation; Ryan Hird, November 2021; https://www.geekymedics.com

[3] A Survey of Convolutional Neural Networks: Analysis, Applications, and Prospects Zewen Li, Fan Liu, Member, IEEE, Wenjie Yang, Shouheng Peng, and Jun Zhou, Senior Member, IEEE

[4] Convolutional neural networks: An overview and application in radiology Rikiya Yamashita, Mizuho Nishio, Richard Kinh Gian Do, Kaori Togashi Received: 3 March 2018 / Revised: 24 April 2018 / Accepted: 28 May 2018 / Published online: 22 June 2018

[5] A Robust Approach for Brain Tumor Detection in Magnetic Resonance Images (Using Fine-tuned EfficientNet) Hasnain Ali Shah, Faisal Saeed, Sangseok Yun, Jun-Hyun Park, Anand Paul and Jae-Mo Kang

[6] Olga Russakovsky, Jia Deng, Hao Su, Jonathan Krause, Sanjeev Satheesh, Sean Ma, Zhiheng Huang, Andrej Karpathy, Aditya Khosla, Michael Bernstein, Alexander C. Berg, & Li Fei-Fei. (2015). ImageNet Large Scale Visual Recognition Challenge.

[7] Kaggle Dataset: https://www.kaggle.com/datasets/navoneel/brain-mri-images-for-brain-tumor-detection?datasetId=165566

[8] Mingxing Tan, & Quoc V. Le. (2020). EfficientNet: Rethinking Model Scaling for Convolutional Neural Networks.

[9] Mark Sandler, Andrew Howard, Menglong Zhu, Andrey Zhmoginov, & Liang-Chieh Chen. (2019). MobileNetV2: Inverted Residuals and Linear Bottlenecks.

[10] Alexey Dosovitskiy, Lucas Beyer, Alexander Kolesnikov, Dirk Weissenborn, Xiaohua Zhai, Thomas Unterthiner, Mostafa Dehghani, Matthias Minderer, Georg Heigold, Sylvain Gelly, Jakob Uszkoreit, & Neil Houlsby. (2021). An Image is Worth 16×16 Words: Transformers for Image Recognition at Scale.

[11] Christian Szegedy, Sergey Ioffe, Vincent Vanhoucke, & Alex Alemi. (2016). Inception-v4, Inception-ResNet and the Impact of Residual Connections on Learning.

[12] Christian Szegedy, Vincent Vanhoucke, Sergey Ioffe, Jonathon Shlens, & Zbigniew Wojna. (2015). Rethinking the Inception Architecture for Computer Vision.

[13] Kaiming He, Xiangyu Zhang, Shaoqing Ren, & Jian Sun. (2015). Deep Residual Learning for Image Recognition.

[14] François Chollet. (2017). Xception: Deep Learning with Depthwise Separable Convolutions.

[15] Prakasa, E. (2016). Texture Feature Extraction by Using Local Binary Pattern. Journal INKOM, 9, 45.

[16] Chaudhari, Akshay; Chambon, Pierre; et al. (2022). RoentGen: Vision-Language Foundation Model for Chest X-Ray Generation (hosted on ArXiv)

